# Burden of COVID-19 pandemic in India: Perspectives from Health Infrastructure

**DOI:** 10.1101/2020.05.26.20113456

**Authors:** Harihar Sahoo, Chaitali Mandal, Suyash Mishra, Snigdha Banerjee

## Abstract

The coronavirus (COVID-19) is spreading rapidly across the country but India’s testing regime is far from the global standards. It is important to identify the states where testing needs expansion and the magnitudes of active COVID cases are higher focusing on current health infrastructure to meet the pandemic. The data on COVID-19 was extracted from the Application Programming Interface. Test positive rate, test per confirmed case, recovery rate, case fatality rate, and percent distribution of active cases were computed. Availability of hospitals, hospital beds, intensive care unit and ventilators per lakh population was also computed by public and private sector. The result revealed that, Maharashtra constitutes more than one-third positive cases in the country. More than a quarter of the active cases in India belonged to the Mumbai district of Maharashtra, followed by the Chennai district (9.4%) and Ahmedabad district (9.1%). Further, about 40 percent of the active cases in India belonged to the 11 districts of Maharashtra. The increased test positive rate in Maharashtra and Gujarat to almost double in last one month is a concern. In order to bring the states and the country in right track, the test positive rate need to be brought down to below 2 percent. The procurement of higher number of high throughput machine, the Cobas 6800 testing machine, is need of the hour. Only few states have adequate health infrastructure. The priority should be the laid on expansion of more laboratories and hospitals, storage of PPE kit, testing kit, and indigenously developed vaccines.

**Highlights:**

- Maharashtra is having the highest number of positive cases followed by Gujarat and Tamil Nadu. Maharashtra constitutes more than one-third positive cases in the country, but the test per confirmed cases (8) is much lower than the other states.
- More than a quarter of the active cases in India belonged to the Mumbai district (26.1%) of Maharashtra, followed by the Chennai district (9.4%) and Ahmedabad district (9.1%). Further, about 40 percent of the active cases in India belonged to the 11 districts of Maharashtra.
- The test positive rate is higher in Maharashtra, Gujarat and Delhi is a concern.
- The recovery rate in India increased substantially by 26.5 percent point from 11.9 percent on April 14 to 38.4 percent on May 17, 2020.
- The case fatality rate of Covid-19 in India declined by 0.2 percent from 3.4 percent on April 14 to 3.2 percent on May 17 in India.
- The number of Dedicated Covid Hospitals is not sufficient in India.
- The available ventilators in the country will deficit in near future to cater to a growing number of active Covid-19 patients and the burden of other communicable and non-communicable diseases.
- India has only 569 testing laboratories (396 govt. and 173 private) against its 1.35 billion population. The procurement of higher number of high throughput machine, the Cobas 6800 testing machine, is need of the hour.

## Genesis

The coronavirus (COVID-19) is spreading rapidly, and scientists are endeavoring to discover drugs for its efficacious treatment in the world (Gao et al., 2020). Covid-19 is an infectious disease; mostly, infected people will experience mild to moderate respiratory illness, high fever, sore throat, nasal congestion, malaise, headache, muscle pain, (Cascella et al., 2020). Older people, suffering some medical problems like cardiovascular disease, diabetes, chronic respiratory disease, and cancer, are more likely to develop severe illness (Remuzzi & Remuzzi, 2020; Singhal, 2020). Currently, there are no specific vaccines or drugs for Covid-19 (Singh et al., 2020). Studies suggest that the use of isolation is the best way to contain this epidemic (Cascella et al., 2020). Thus, countries are racing to reduce the spread of the virus by treating and testing patients, limiting travel, carrying out contact tracing, quarantining citizens, and cancelling large gatherings such as sporting events, concerts, and schools (United Nation Development Programme).

The Covid-19 disease surfaced first in December, 2019, which is linked to direct exposure to the Huanan seafood wholesale market of Wuhan, China (Velavan, & Meyer, 2020). On January 30, the World Health Organisation (WHO) declared this Covid-19 situation as a Public Health Emergency of International Concern (Kannan et al. 2020). India has reported 95676 confirmed Covid-19 cases on May 17, 2020. India closed its international borders and enforced an immediate lockdown on March 24, 2020, for 21 days and it further extended till May 31, 2020, which WHO praised as ‘tough and timely’. The government demarcated 130 districts as red zones (Hotspot area), 284 districts as orange zones, and 319 districts as green zones, on May 1, 2020. The list identifies all 11 regions of Delhi as red zones. Other metropolitan cities, including Mumbai, Bengaluru, Hyderabad, Chennai, and Kolkata have also been designated as red zones (Nair, 2020).

As the number of Covid-19 positive case is increasing day by day, the study on test rate is important. If a country has fewer cases than expected, it could be either because the country did a good job to combat against the virus or the country is not testing for the virus well enough. The testing facility is not the same across the states and union territories. Rajasthan, the largest state in terms of the area, has the same number (eight) of testing labs as the city of Chennai, and that is just double India’s most densely populated state Bihar. Maharashtra has the highest testing centres and the maximum number of Covid-19 cases, on the other in the spectrum, Nagaland has no testing centre and no reported case (Banerjee, 2020). West Bengal experienced the highest death rate from Covid-19, along with the lower testing than major states in India (Basu, 2020b). The reported cases (confirmed, active, recovery and death) differ across the states because of the wide variation in the testing centres. The number of testing indicates, some states are farther behind in getting a sense of the outbreak than others (Chauhan and Kawoosa, 2020). India’s testing regime is far from the global standards (Basu, 2020a).

The state governments began taking more swab samples, without realizing the limited capacity of laboratories in the states. Many states have sent samples to laboratories outside the state to get faster test results (Thakur et al., 2020). That is why there is a need to study the current trend in the testing, recovery rates, and fatality rates. Study on active case is important as they really need health care. Because of geographical and socio-economic diversity in the country, there is a wide variation in health infrastructure. Preparedness and response to Covid-19 is not same at the state level. Health inequalities, widening economic and social disparities, and distinct cultural values in India, present unique challenges. In India, however, the public health system already overburdened with various Communicable and Non-communicable diseases. The implementation of the public health measures to handle the epidemic is difficult in places with inadequate hygiene and sanitation and overcrowded living conditions. Thus the study on current health infrastructure is important to meet the pandemic. With this background, the specific objectives of the study are as follows:

1. To identify the states where the testing need to be expanded.
2. To study the change in recovery and fatality rate and to identify the areas where the magnitude of active Covid cases are higher.
3. To link the availability of health infrastructure with Covid-19 positive cases.

### Data and Methods

The Covid-19 related information is taken from the Application Programming Interface (API, https://api.Covid19india.org/csv/) a data-sharing portal that provides the most updated information on the daily and total confirmed cases, active cases, recovered cases, and deaths for each affected states/Union Territories. The data on number of government hospital and hospital beds are obtained from National Health Profile (2019). The estimated number of private hospital, private hospital beds and intensive care unit (ICU) beds and ventilators are taken from a report of The Center for Disease Dynamics, Economics & Policy (Kapoor et al., 2020). The projected population of 2020 data is obtained from the Report of the Technical Group on Population Projection (RGI, 2019).

The analysis in the study is performed upto the end of Lockdown 3, i.e., 17^th^ May, 2020. The phases were upto the end of 1^st^ Lockdown i.e., 14^th^ April 2020; upto the 2^nd^ Lockdown (3^rd^ May 2020); and finally upto the 3^rd^ Lockdown (17^th^ May 17 2020). The test positivity rate is defined as the proportion positive cases per 100 persons tested. Test per confirmed case is defined as the number of tests performed per positive case. Tests per million population are also calculated. Further, recovery rate, case fatality rate, active rate and percent distribution of active cases of Covid-19 in different phases are also computed. The hotspot zones of confirmed positive cases in the districts are identified using the GIS software. Availability of hospitals, hospital beds, intensive care unit (ICU) beds and ventilators per lakh population is also computed by public, private and total.

## Results

### Covid-19 Testing in India and states

**Table 1** presents the scenario of Covid-19 testing till 17^th^ May, 2020, for states of India. Results show that, till 17^th^ May, a total of 2457651 individuals were tested for COVID-19 in India, out of which 95676 individuals were tested positive. The total number of active cases as on 17^th^ May was reported as 55868. Tamil Nadu followed by Andhra Pradesh, Maharashtra, Rajasthan and Uttar Pradesh recorded the highest number of individuals tested for COVID-19 with Maharashtra having the highest number of positive cases followed by Gujarat and Tamil Nadu. It is clear that, in India, about 1824 individuals were tested per million populations varying from 377 individuals in Bihar to 6725 in Delhi. Further, Maharashtra, followed by Tamil Nadu, Gujarat, Delhi, and Madhya Pradesh, were the states with the highest number of active cases as on 17^th^ May. The test positivity rate in India was 3.9 percent, and it varied largely across the states of India, varying from 12.1 percent in Maharashtra to 0.7 percent in Jharkhand. The test positivity rate was higher than the national average in six states, namely Maharashtra (12.1%), Gujarat (7.9%), Delhi (7.2%), Chandigarh (6.8%), Telangana (6.6%) and Madhya Pradesh (4.8%). The test per confirmed cases in India was estimated as 26, and wide variation was observed across the states varying from 8 in Maharashtra to 149 in Jharkhand. While the test per confirmed cases in most of the states was higher than the national average, it was lower in six states namely Maharashtra (8), Gujarat (14), Delhi (13), Chandigarh (15), Telangana (15) and Madhya Pradesh (21). Thus looking at the testing and positive cases, states, namely Kerala, Karnataka, Andhra Pradesh, Odisha, Jharkhand, and Haryana, are doing better than the other states in terms of testing Covid-19. On the other hand, states which have a higher proportion of positive cases, i.e., Maharashtra, Gujarat, Delhi, and Madhya Pradesh, testing seems to be much lower. Maharashtra constitutes more than one-third positive cases in the country, but the test per confirmed cases (8) is much lower than the other states. Similarly, Gujarat and Delhi each constitute little more than 10 percent of positive cases in the country, but the test per confirmed cases is 13 and 14, respectively. Thus it clearly indicates that there is an urgent need to increase the Covid-19 testing centers in these states.

**Table 1:** Test Positive Rate, Tests per confirmed case, Tests per million population by State, India as on May 17 2020

| State | Projected<br>Population (in 000) as<br>on March 31 2020 | Total<br>Tested | Total<br>Positive<br>Cases | Recovered<br>Cases | Deceased<br>Cases | Active<br>Cases | Test<br>positivity<br>rate | Tests Per<br>Confirmed<br>Case | Tests per<br>million<br>population |
| --- | --- | --- | --- | --- | --- | --- | --- | --- | --- |
| Maharashtra | 123295 | 274040 | 33053 | 7688 | 1197 | 24168 | 12.1 | 8 | 2223 |
| Tamil Nadu | 76049 | 326720 | 11224 | 4172 | 79 | 6973 | 3.4 | 29 | 4296 |
| Gujarat | 68862 | 143600 | 11380 | 4499 | 659 | 6222 | 7.9 | 13 | 2085 |
| Delhi | 20193 | 135791 | 9755 | 4202 | 148 | 5405 | 7.2 | 14 | 6725 |
| Madhya Pradesh | 83374 | 103898 | 4977 | 2403 | 249 | 2325 | 4.8 | 21 | 1246 |
| Rajasthan | 78273 | 231946 | 5202 | 3045 | 131 | 2026 | 2.2 | 45 | 2963 |
| Uttar Pradesh | 227943 | 172219 | 4464 | 2636 | 112 | 1716 | 2.6 | 39 | 756 |
| West Bengal | 97516 | 85956 | 2677 | 959 | 238 | 1480 | 3.1 | 32 | 881 |
| Andhra Pradesh | 52504 | 238998 | 2362 | 1456 | 50 | 856 | 1.0 | 101 | 4552 |
| Bihar | 121302 | 45729 | 1320 | 473 | 8 | 839 | 2.9 | 35 | 377 |
| Odisha | 43852 | 91223 | 828 | 220 | 4 | 604 | 0.9 | 110 | 2080 |
| Karnataka | 66322 | 145398 | 1147 | 509 | 37 | 601 | 0.8 | 127 | 2192 |
| Jammu and Kashmir | 13305 | 80934 | 1183 | 575 | 13 | 595 | 1.5 | 68 | 6083 |
| Punjab | 30099 | 51812 | 1964 | 1366 | 35 | 563 | 3.8 | 26 | 1721 |
| Telangana | 37473 | 23388 | 1551 | 992 | 34 | 525 | 6.6 | 15 | 624 |
| Haryana | 29077 | 78029 | 910 | 561 | 14 | 335 | 1.2 | 86 | 2684 |
| Chandigarh | 1193 | 2812 | 191 | 51 | 3 | 137 | 6.8 | 15 | 2357 |
| Jharkhand | 37937 | 33220 | 223 | 113 | 3 | 107 | 0.7 | 149 | 876 |
| Kerala | 35307 | 45027 | 602 | 497 | 4 | 101 | 1.3 | 75 | 1275 |
| Tripura | 4032 | 13750 | 164 | 85 | 0 | 79 | 1.2 | 84 | 3410 |
| Rest of India | 99826 | 133161 | 500 | 283 | 6 | 211 | 0.4 | 266 | 1334 |
| **India** | **1347734** | **2457651** | **95676** | **36785** | **3023** | **55868** | **3.9** | **26** | **1824** |

The test positivity rate in India varied from 4.7 percent on April 14 to 3.9 percent on May 17 (**Figure 1**). A large variation in test positivity rate was observed across the selected states of India. While the test positivity rate has increased in the states of Maharashtra, Gujarat, and Punjab during this period, the rate declined in case of remaining states. The increase in test positivity rate was highest in Maharashtra (6.4%) followed by Gujarat (3.8%) whereas the decline was highest in Madhya Pradesh (4.3%) followed by Andhra Pradesh (3.5%), West Bengal (3.1%) and Tamil Nadu (2.9%). The decline in test positivity rate remained low in case Uttar Pradesh (0.7%), Rajasthan (0.6%), and Delhi (0.6%). In India, the test per confirmed cases increased by five individuals from 21 on April 14 to 26 on May 17 (**Figure 2**). The test per confirmed cases declined in the case of Maharashtra and Gujarat while it remained the same in the case of Punjab. While the test per confirmed cases declined by seven individuals in Maharashtra and four individuals in Gujarat, it increased by 75 individuals in Andhra Pradesh, 15 individuals in West Bengal, 13 individuals in Tami Nadu, ten individuals in Madhya Pradesh and Rajasthan, six individuals in Uttar Pradesh and four individuals in Delhi.

**Figure 1:**
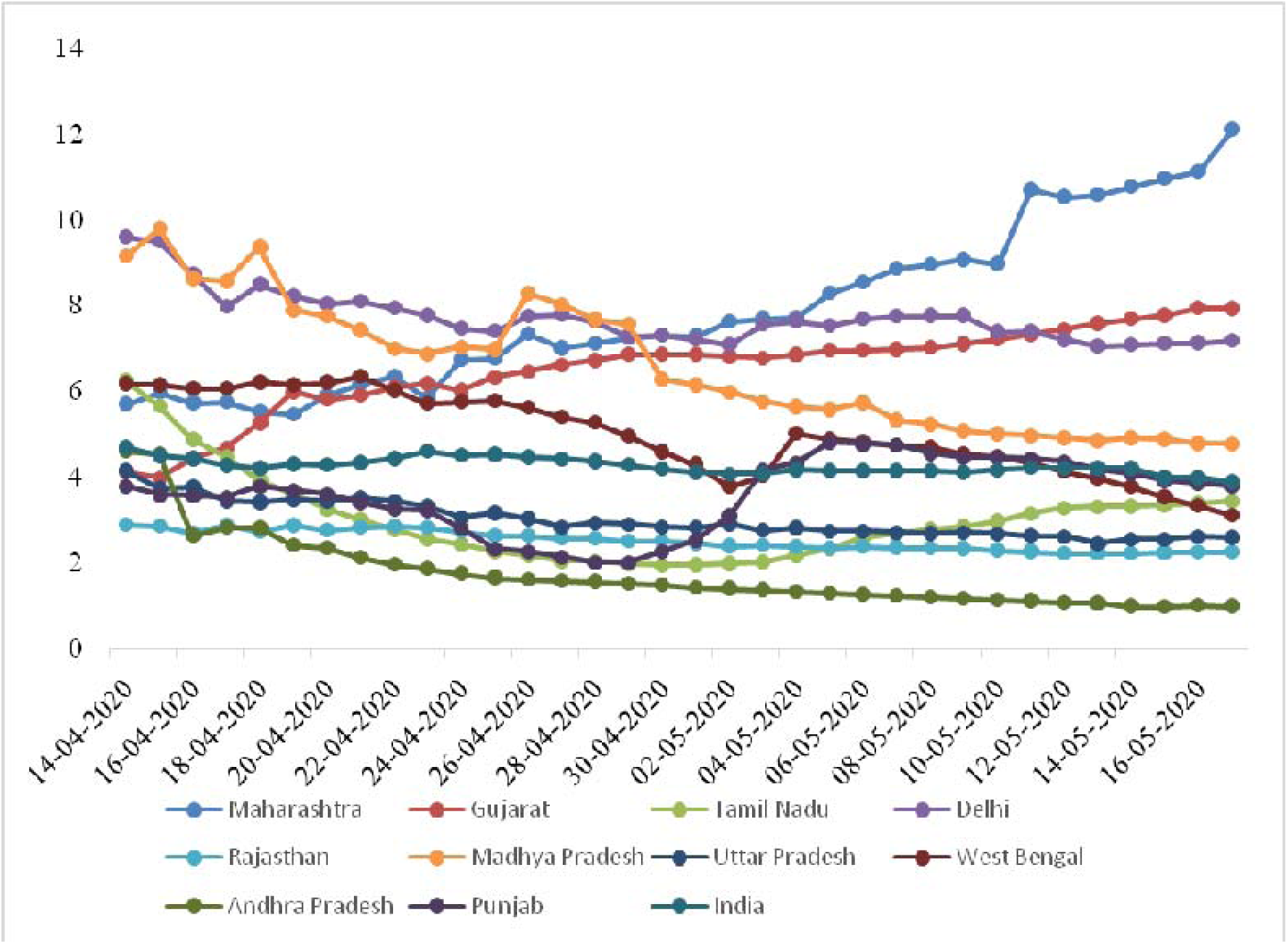
Change in the Test Positive rate of COVID-19

**Figure 2:**
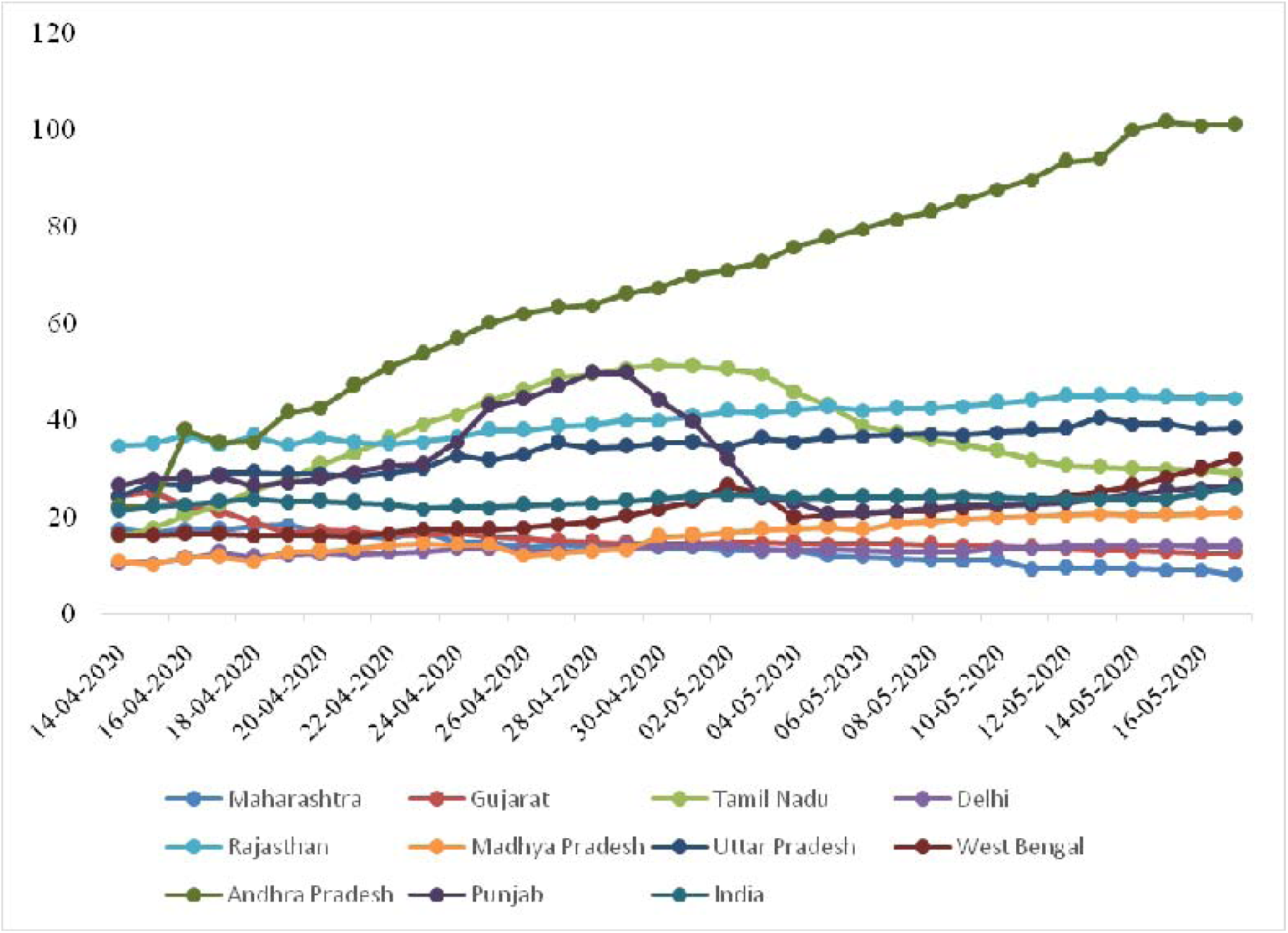
Change in Tests per Confirmed Case of COVID-19

### Change in recovery, fatality rate, and the hotspot areas

The recovery rate in India is increased substantially by 26.5 percent point from 11.9 percent on April 14 to 38.4 percent on May 17 (**Figure 3**). All the selected states exhibited an increase in recovery rate during this period; however, wide variation across states was observed. Andhra Pradesh, Punjab, and Uttar Pradesh showed a change in recovery rate of more than fifty percent. The change in recovery rate was highest in Andhra Pradesh (58.3%), followed by Punjab (54.9%), Tripura (51.8%), Uttar Pradesh (51.5%), Rajasthan (43.9%), Delhi (41.1%) and Madhya Pradesh (39.6%) while the changes were lowest in case Maharashtra (12.2%) followed by West Bengal (16.9%), Gujarat (30 %) and Tamil Nadu (30.5%).

**Figure 3:**
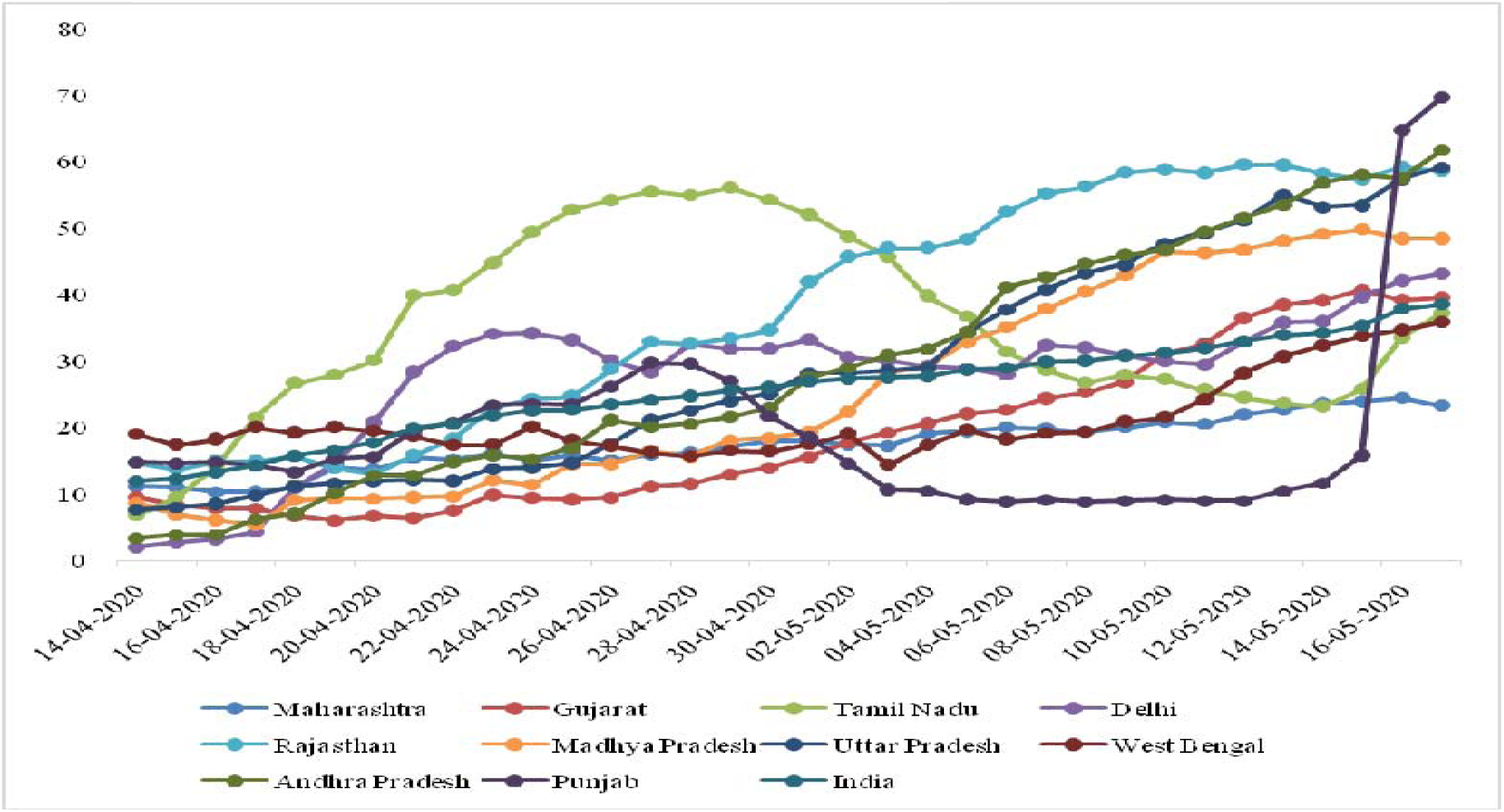
Change in recovery rate of COVID-19 by most affected states of India

The case fatality rate of Covid-19 in India has declined by 0.2 percent from 3.4 percent on April 14 to 3.2 percent on May 17 in India (**Figure 4**). While the change in case fatality rate was negative in the state of Punjab, Maharashtra, Tamil Nadu, Madhya Pradesh, Andhra Pradesh, and Delhi, it was positive in case of West Bengal, Rajasthan, Gujarat, and Uttar Pradesh. The negative change in case fatality rate highest in the case of Punjab (5.3%), followed by Maharashtra (3.9%) and Madhya Pradesh (2.3%). In contrast, the positive change in case fatality rate was highest in West Bengal (5.2%) followed by Rajasthan (1.4%), Gujarat (1.3%) and Uttar Pradesh (1.3%).

**Figure 4:**
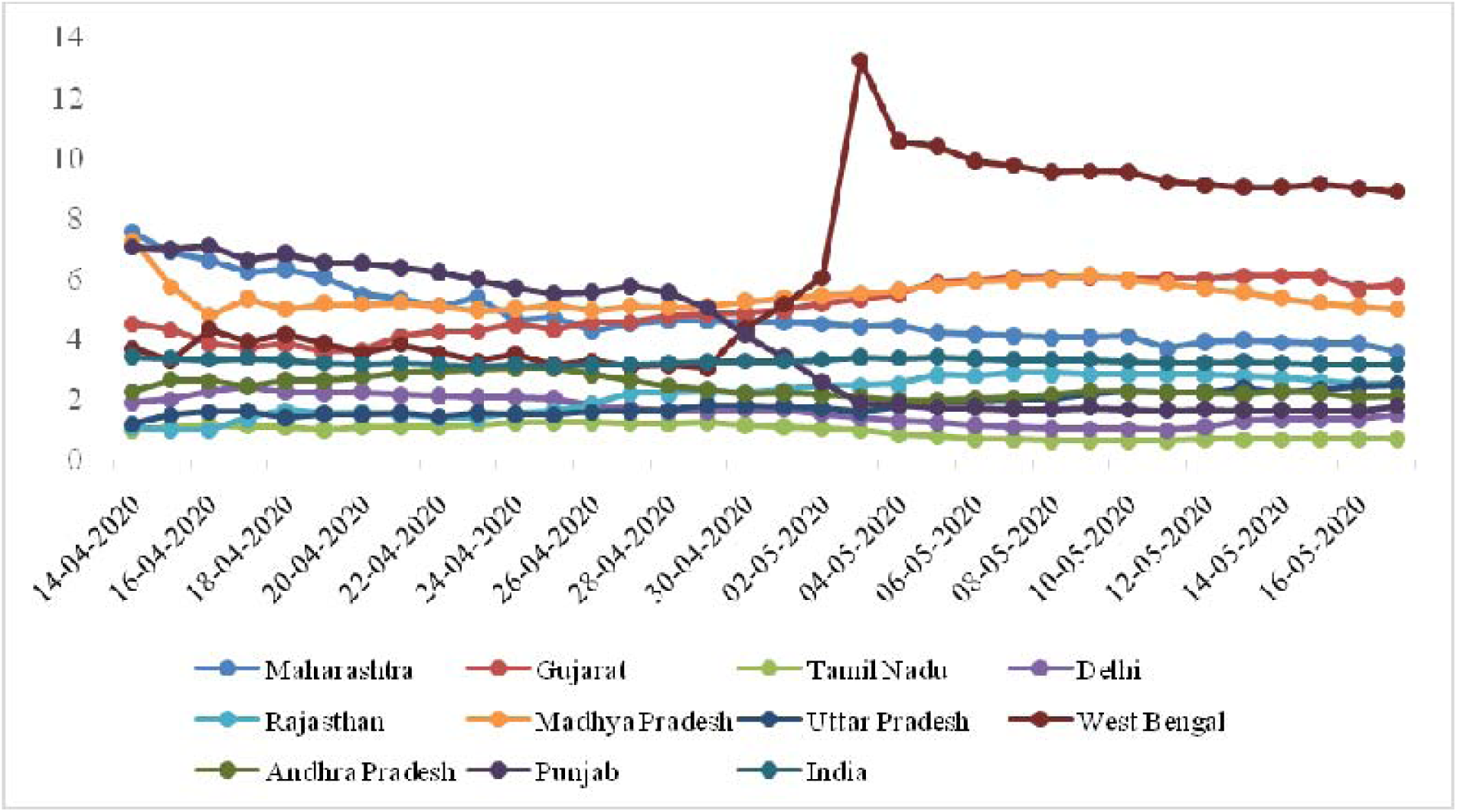
Change in fatality rate of COVID-19 by most affected states of India

The recovery rate of Covid-19 in India was 38.4, while the case fatality rate was 3.2 percent and exhibited wide variation across selected states (**Table 2**). The recovery rate was highest in Kerala (82.6%), followed by Punjab, Telangana, Andhra Pradesh, and Haryana, while it was more than fifty percent in the states of Uttar Pradesh, Rajasthan, Tripura, and Jharkhand. Further, the recovery rate was lowest in Maharashtra (23.1%), followed by Odisha, Chandigarh. The case fatality of Covid-19 was highest in West Bengal (8.9%), followed by Gujarat (5.8%) and Madhya Pradesh (5%) while it was lowest in Tripura, Odisha, Bihar, Tamil Nadu, and Kerala. More than three-quarters of the active cases belonged to the state of Maharashtra (4.3.3%), Tamil Nadu (12.5%), Gujarat (11.1), and Delhi (9.7%).

**Table 2:**
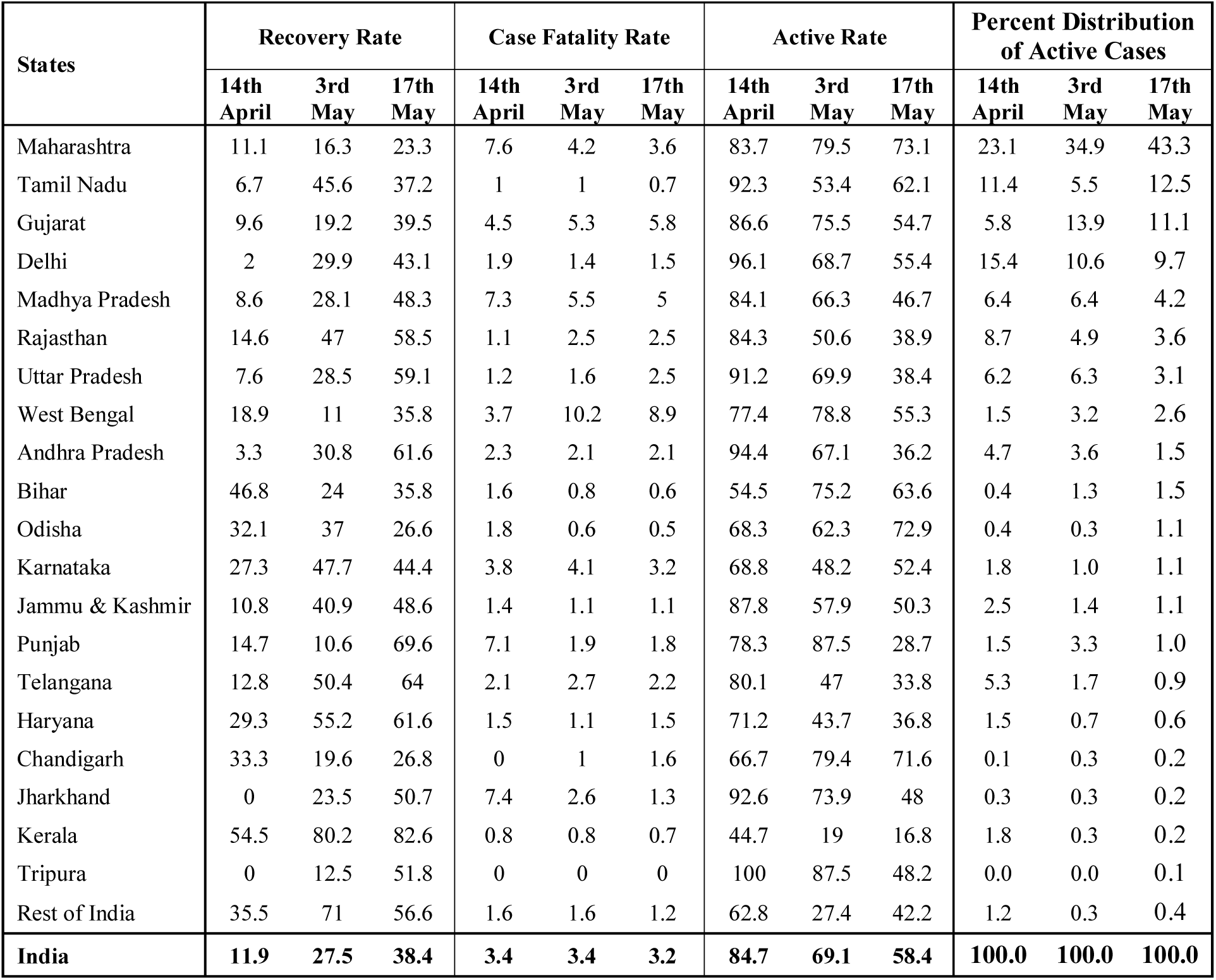
Change in the Recovery Rate, Case Fatality Rate and Active Rate by State, India

| States | Recovery Rate |  |  | Case Fatality Rate |  |  | Active Rate |  |  | Percent Distribution of Active Cases |  |  |
| --- | --- | --- | --- | --- | --- | --- | --- | --- | --- | --- | --- | --- |
|  | 14th April | 3rd May | 17th May | 14th April | 3rd May | 17th May | 14th April | 3rd May | 17th May | 14th April | 3rd May | 17th May |
| Maharashtra | 11.1 | 16.3 | 23.3 | 7.6 | 4.2 | 3.6 | 83.7 | 79.5 | 73.1 | 23.1 | 34.9 | 43.3 |
| Tamil Nadu | 6.7 | 45.6 | 37.2 | 1 | 1 | 0.7 | 92.3 | 53.4 | 62.1 | 11.4 | 5.5 | 12.5 |
| Gujarat | 9.6 | 19.2 | 39.5 | 4.5 | 5.3 | 5.8 | 86.6 | 75.5 | 54.7 | 5.8 | 13.9 | 11.1 |
| Delhi | 2 | 29.9 | 43.1 | 1.9 | 1.4 | 1.5 | 96.1 | 68.7 | 55.4 | 15.4 | 10.6 | 9.7 |
| Madhya Pradesh | 8.6 | 28.1 | 48.3 | 7.3 | 5.5 | 5 | 84.1 | 66.3 | 46.7 | 6.4 | 6.4 | 4.2 |
| Rajasthan | 14.6 | 47 | 58.5 | 1.1 | 2.5 | 2.5 | 84.3 | 50.6 | 38.9 | 8.7 | 4.9 | 3.6 |
| Uttar Pradesh | 7.6 | 28.5 | 59.1 | 1.2 | 1.6 | 2.5 | 91.2 | 69.9 | 38.4 | 6.2 | 6.3 | 3.1 |
| West Bengal | 18.9 | 11 | 35.8 | 3.7 | 10.2 | 8.9 | 77.4 | 78.8 | 55.3 | 1.5 | 3.2 | 2.6 |
| Andhra Pradesh | 3.3 | 30.8 | 61.6 | 2.3 | 2.1 | 2.1 | 94.4 | 67.1 | 36.2 | 4.7 | 3.6 | 1.5 |
| Bihar | 46.8 | 24 | 35.8 | 1.6 | 0.8 | 0.6 | 54.5 | 75.2 | 63.6 | 0.4 | 1.3 | 1.5 |
| Odisha | 32.1 | 37 | 26.6 | 1.8 | 0.6 | 0.5 | 68.3 | 62.3 | 72.9 | 0.4 | 0.3 | 1.1 |
| Karnataka | 27.3 | 47.7 | 44.4 | 3.8 | 4.1 | 3.2 | 68.8 | 48.2 | 52.4 | 1.8 | 1.0 | 1.1 |
| Jammu & Kashmir | 10.8 | 40.9 | 48.6 | 1.4 | 1.1 | 1.1 | 87.8 | 57.9 | 50.3 | 2.5 | 1.4 | 1.1 |
| Punjab | 14.7 | 10.6 | 69.6 | 7.1 | 1.9 | 1.8 | 78.3 | 87.5 | 28.7 | 1.5 | 3.3 | 1.0 |
| Telangana | 12.8 | 50.4 | 64 | 2.1 | 2.7 | 2.2 | 80.1 | 47 | 33.8 | 5.3 | 1.7 | 0.9 |
| Haryana | 29.3 | 55.2 | 61.6 | 1.5 | 1.1 | 1.5 | 71.2 | 43.7 | 36.8 | 1.5 | 0.7 | 0.6 |
| Chandigarh | 33.3 | 19.6 | 26.8 | 0 | 1 | 1.6 | 66.7 | 79.4 | 71.6 | 0.1 | 0.3 | 0.2 |
| Jharkhand | 0 | 23.5 | 50.7 | 7.4 | 2.6 | 1.3 | 92.6 | 73.9 | 48 | 0.3 | 0.3 | 0.2 |
| Kerala | 54.5 | 80.2 | 82.6 | 0.8 | 0.8 | 0.7 | 44.7 | 19 | 16.8 | 1.8 | 0.3 | 0.2 |
| Tripura | 0 | 12.5 | 51.8 | 0 | 0 | 0 | 100 | 87.5 | 48.2 | 0.0 | 0.0 | 0.1 |
| Rest of India | 35.5 | 71 | 56.6 | 1.6 | 1.6 | 1.2 | 62.8 | 27.4 | 42.2 | 1.2 | 0.3 | 0.4 |
| **India** | **11.9** | **27.5** | **38.4** | **3.4** | **3.4** | **3.2** | **84.7** | **69.1** | **58.4** | **100.0** | **100.0** | **100.0** |

The recovery rate in India by the end of 1^st^ lockdown was 11.9 percent, which increased to 27.5 percent at the end of 2^nd^ lockdown and 38.4 percent by the end of 3^rd^ lockdown showing a positive change of 15.6 percent during 2^nd^ lockdown phase and 10.9 percent during 3^rd^ lockdown phase. On the contrary, the case fatality rate showed no change during the 2^nd^ lockdown phase while negative change of 0.2 percent point by the end of 3^rd^ lockdown phase. Similarly, the proportion of active cases has declined by 15.6 percent and 10.9 percent by the end of second and third lockdown, respectively. The recovery rate was highest in Tamil Nadu (38.9%), followed by Telangana (35.5%) and Rajasthan (33.3%), while it was lowest in Maharashtra (6.6%) followed by Odisha (7.0%) and Gujarat (10.1%) during the 2^nd^ lockdown phase. Further, negative growth in recovery rate was observed in Bihar (19.9%), Chandigarh (13.7%), West Bengal (7.9%) and Punjab (4.1%). By the end of the 3^rd^ phase of lockdown, Punjab (59%) followed by Tripura (39.3%) and Andhra Pradesh (30.8%) recorded the higher recovery rate while Kerala (2.4%) followed by Haryana (6.4%) and Maharashtra (7%) had the lower recovery rate. During a similar phase, negative growth in recovery rate was observed in Odisha (10.4%), Tamil Nadu (8.4%) and Karnataka (3.3%). In case of case fatality rate, during 2^nd^ lockdown phase, West Bengal (6.5%) followed by Rajasthan (1.4%), Gujarat (1%), Chandigarh (1%), Uttar Pradesh (0.4%) and Karnataka (0.3%) had positive case fatality rate while negative or no growth was observed in remaining states. Punjab (5.2%) recorded the highest decline in case fatality rate followed by Jharkhand (4.8%) and Maharashtra (2.4%). During 3^rd^ phase of lockdown, while the state of Uttar Pradesh (0.9%), Chandigarh (0.6%), Gujarat (0.5%), Haryana (0.4%) and Delhi (0.1%) recorded positive growth in case fatality rate, negative or no growth was observed in the remaining states with Jharkhand (1.3%) followed by West Bengal (1.3%) and Maharashtra (0.9%) recording the highest negative growth. During 2^nd^ phase of lockdown, the active number of cases increased by 20.7 percent in Bihar followed by Chandigarh (12.7%), Punjab (9.2%) and West Bengal (1.4%), negative growth was observed in remaining states with the highest decline in active cases observed in Tamil Nadu (38.9%) followed by Rajasthan (33.7%) and Telangana (33.1%). During the 3^rd^ phase, while Odisha, Tamil Nadu, and Karnataka observed a surge in the active number of cases, remaining states observed a decline in active cases. The decline in active rate was highest in Punjab (58.8%), followed by Tripura (39.3%) and Uttar Pradesh (31.5%).

In case of percent distribution of active cases, during 2^nd^ phase of lockdown in India, Maharashtra followed by Gujarat, Punjab, West Bengal, Bihar, Chandigarh, and Uttar Pradesh recorded a positive change in the distribution of active cases whereas negative or no growth was observed in the distribution of active cases in the remaining states. The decline in the distribution of active cases was highest in Tamil Nadu (6%), followed by Delhi (4.9%) and Rajasthan (3.8%). Similarly, during 3^rd^ phase of lockdown, the change in the distribution of active cases was positive in Maharashtra, followed by Tamil Nadu, Odisha, Bihar, Tripura, and Karnataka, while negative or zero change in the distribution of active cases was observed in the remaining states. The decline in the distribution of active cases was highest in Uttar Pradesh (3.2%), followed by Gujarat (2.7%) and Punjab (2.3).

### High Focused Districts in India

**Table 3** presents the scenario of recovery rate and case fatality rate of the top 40 high focused district of India based on active cases of Covid-19 till May 17, 2020. The top 40 districts accounted for 64.1 percent of the total confirmed cases in India, 76.9 percent of the total deceased cases, and 72.8 percent of the total active cases. The average recovery rate of these forty districts was estimated as 30 percent, while the case fatality rate was 3.8 percent, and wide variation was observed across the districts. Of the top 40 districts, 11 districts belonged to Maharashtra, five districts from Tamil Nadu, four districts from Madhya Pradesh, three districts each from Gujarat, Rajasthan and West Bengal, two districts each from Delhi, Uttar Pradesh, Andhra Pradesh and Jammu & Kashmir and one district each from Odisha, Telangana, and Chandigarh. The recovery rate was highest in the Jodhpur district (71.9%) of Rajasthan followed by Kurnool (66.3%) and Guntur district (65.9%) of Andhra Pradesh and Surat district (65.5%) of Gujarat while it was lowest in South East (0.0%) and Central district (0.0%) of Delhi followed by Udaipur district (3.2%) of Rajasthan and Kulgam district (6.1%) of Jammu & Kashmir. The case fatality rate was highest in Ujjain district (14.3%) of Madhya Pradesh followed by Kolkata (12.2%) and North 24 Parganas districts of West Bengal and Burhanpur district of Madhya Pradesh while it was lowest in Kulgam district (0%) of Jammu & Kashmir, Udaipur district (0%) of Rajasthan, South East (0%) and Central district (0%) of Delhi. Ahmedabad district of Gujarat accounted for 84.3 percent of the active cases in Gujarat, while the Chennai district (75.2%) of Tamil Nadu accounted for three-quarters of the active cases in the states. About 65 percent of active cases in the state of Maharashtra belonged to the Mumbai district, whereas Indore district (53.8%) of Madhya Pradesh accounted for more than half of the active cases in the state. More than a quarter of the active cases in India belonged to the Mumbai district (26.1%) of Maharashtra, followed by the Chennai district (9.4%) of Tamil Nadu and Ahmedabad district (9.1%) of Gujarat. Further, about 40 percent of the active cases in India belonged to the 11 districts of Maharashtra.

**Table 3:** High Focused districts (Top 40 districts) in active cases as on May 17 2020

| Sl. No. | District | State | Confirmed Cases | Recovered Cases | Deceased Cases | Active Cases | Recovery Rate | Case Fatality Rate | Percent Active Cases within State | Percent Active Cases within India |
| --- | --- | --- | --- | --- | --- | --- | --- | --- | --- | --- |
| 1 | Mumbai | Maharasht | 18555 | 3260 | 696 | 1459 | 17.6 | 3.8 | 64.9 | 26.13 |
| 2 | Chennai | Tamil | 6761 | 1476 | 54 | 5231 |  | 0.8 | 75.2 | 9.36 |
| 3 | Ahmedab | Gujarat | 8144 | 2545 | 493 | 5106 | 31.3 | 6.1 | 84.3 | 9.14 |
| 4 | Thane | Maharasht | 3834 | 766 | 47 | 3021 | 20.0 | 1.2 | 13.4 | 5.41 |
| 5 | Pune | Maharasht | 3647 | 1452 | 188 | 2007 | 39.8 | 5.2 | 8.9 | 3.59 |
| 6 | Indore | Madhya | 2470 | 1119 | 100 | 1251 | 45.3 | 4.0 | 53.8 | 2.24 |
| 7 | Kolkata | West | 1311 | 478 | 160 | 673 | 36.5 | 12.2 | 46.0 | 1.20 |
| 8 | Hyderaba | Telangana | 981 | 305 | 23 | 653 | 31.1 | 2.3 | 74.6 | 1.17 |
| 9 | Aurangab | Maharasht | 873 | 260 | 25 | 588 | 29.8 | 2.9 | 2.6 | 1.05 |
| 10 | Jaipur | Rajasthan | 1553 | 901 | 66 | 586 | 58.0 | 4.2 | 29.9 | 1.05 |
| 11 | Nashik | Maharasht | 835 | 265 | 35 | 535 | 31.7 | 4.2 | 2.4 | 0.96 |
| 12 | Bhopal | Madhya | 992 | 564 | 38 | 390 | 56.9 | 3.8 | 16.8 | 0.70 |
| 13 | Udaipur | Rajasthan | 379 | 12 | 0 | 367 | 3.2 | 0.0 | 18.7 | 0.66 |
| 14 | Howrah | West | 571 | 173 | 32 | 366 | 30.3 | 5.6 | 25.0 | 0.66 |
| 15 | Thiruvall | Tamil | 544 | 178 | 5 | 361 | 32.7 | 0.9 | 5.2 | 0.65 |
| 16 | Surat | Gujarat | 1049 | 687 | 49 | 313 | 65.5 | 4.7 | 5.2 | 0.56 |
| 17 | Raigad | Maharasht | 414 | 99 | 12 | 303 | 23.9 | 2.9 | 1.3 | 0.54 |
| 18 | Chengalp | Tamil | 491 | 187 | 3 | 301 | 38.1 | 0.6 | 4.3 | 0.54 |
| 19 | Agra | Uttar | 806 | 501 | 27 | 278 | 62.2 | 3.3 | 16.2 | 0.50 |
| 20 | Jodhpur | Rajasthan | 1004 | 722 | 16 | 266 | 71.9 | 1.6 | 13.6 | 0.48 |
| 21 | Palghar | Maharasht | 390 | 136 | 13 | 241 | 34.9 | 3.3 | 1.1 | 0.43 |
| 22 | Ganjam | Odisha | 292 | 49 | 2 | 241 | 16.8 | 0.7 | 39.9 | 0.43 |
| 23 | Solapur | Maharasht | 371 | 109 | 22 | 240 | 29.4 | 5.9 | 1.1 | 0.43 |
| 24 | Vadodara | Gujarat | 639 | 384 | 32 | 223 | 60.1 | 5.0 | 3.7 | 0.40 |
| 25 | Meerut | Uttar | 322 | 95 | 18 | 209 | 29.5 | 5.6 | 12.2 | 0.37 |
| 26 | Nagpur | Maharasht | 352 | 159 | 2 | 191 | 45.2 | 0.6 | 0.8 | 0.34 |
| 27 | North 24 | West | 355 | 133 | 32 | 190 | 37.5 | 9.0 | 13.0 | 0.34 |
| 28 | Kurnool | Andhra | 611 | 405 | 19 | 187 | 66.3 | 3.1 | 25.0 | 0.33 |
| 29 | Central | Delhi | 184 | 0 | 0 | 184 | 0.0 | 0.0 | 23.4 | 0.33 |
| 30 | Jalgaon | Maharasht | 250 | 52 | 30 | 168 | 20.8 | 12.0 | 0.7 | 0.30 |
| 31 | Cuddalor | Tamil | 417 | 259 | 1 | 157 | 62.1 | 0.2 | 2.3 | 0.28 |
| 32 | Akola | Maharasht | 236 | 71 | 14 | 151 | 30.1 | 5.9 | 0.7 | 0.27 |
| 33 | Anantnag | Jammu & | 177 | 35 | 1 | 141 | 19.8 | 0.6 | 23.7 | 0.25 |
| 34 | Kulgam | Jammu | 147 | 9 | 0 | 138 | 6.1 | 0.0 | 23.2 | 0.25 |
| 35 | Chandiga | Chandigar | 191 | 51 | 3 | 137 | 26.7 | 1.6 | 100.0 | 0.25 |
| 36 | Guntur | Andhra | 417 | 275 | 8 | 134 | 65.9 | 1.9 | 17.9 | 0.24 |
| 37 | Ujjain | Madhya | 329 | 148 | 47 | 134 | 45.0 | 14.3 | 5.8 | 0.24 |
| 38 | South | Delhi | 130 | 0 | 0 | 130 | 0.0 | 0.0 | 16.5 | 0.23 |
| 39 | Tirunelve | Tamil | 195 | 65 | 1 | 129 | 33.3 | 0.5 | 1.9 | 0.23 |
| 40 | Burhanpu | Madhya | 149 | 14 | 10 | 125 | 9.4 | 6.7 | 5.4 | 0.22 |
| **Total** |  |  | **61368** | **18399** | **2324** | **4064** | **30.0** | **3.8** |  | **72.75** |

**Map 1:**
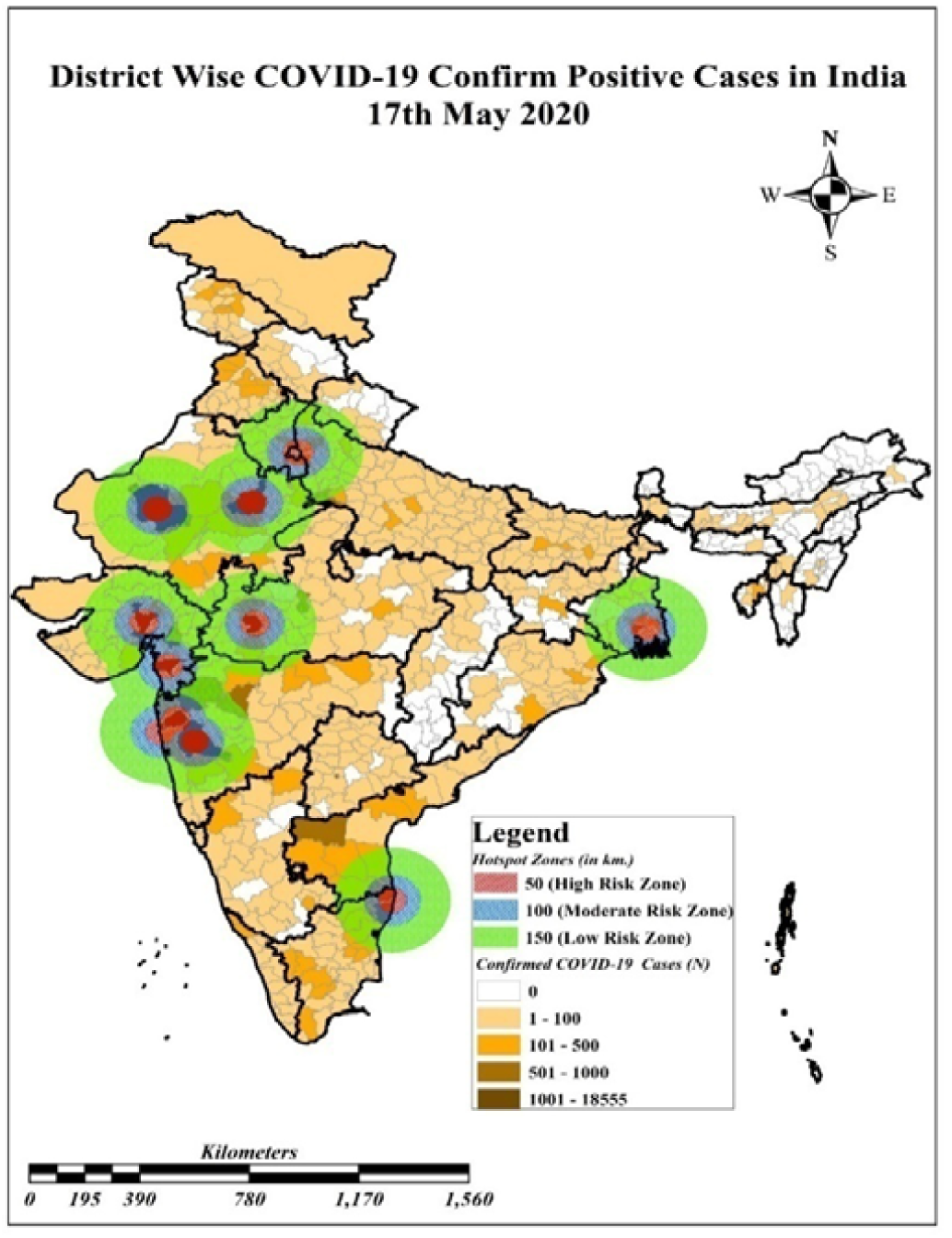
Covid-19 Hotspot zones of India

### Availability of Health infrastructure

**Table 4** represents the availability of health infrastructure in India. There are only 5.1 hospitals per one lakh population in India. The number of government hospitals is less (1.9) compared to the private (3.2) hospitals in the country. India has 140 hospital beds (53 in govt. and 87.9 in private), 7.0 ICU beds (2.6 in govt. and in 4.4 private), and 3.5 ventilators (1.3 in govt. and 2.2 in private) per lakh population. India has not been a great spender on health; it spends only 1.28% of its GDP as public expenditure on health (NHP, 2019). India has just one bed per 1844 patients in government hospitals, and the doctor-patient ratio is 1:1445, far from the WHO recommended one doctor per 1000 patients (Sharma, 2020). Among the states, only a few states have adequate health infrastructure. Karnataka has the highest number of the hospital (16 per lakh population) followed by Telangana, whereas Delhi has recorded the lowest number of hospitals (0.9 per lakh population) followed by Chandigarh, Madhya Pradesh, Jammu and Kashmir, and Andhra Pradesh.

**Table 4:** Availability of health infrastructure per lakh population in India

| States | Number of hospital |  |  | Hospital Beds |  |  | ICU Beds |  |  | Ventilators |  |  |
| --- | --- | --- | --- | --- | --- | --- | --- | --- | --- | --- | --- | --- |
|  | Govt. | Private | Total | Govt. | Private | Total | Govt. | Private | Total | Govt. | Private | Total |
| Maharashtra | 0.6 | 2.0 | 2.6 | 41.7 | 146.2 | 188.0 | 2.1 | 7.3 | 9.4 | 1.0 | 3.7 | 4.7 |
| Tamil Nadu | 1.6 | 1.6 | 3.2 | 102.0 | 102.4 | 204.3 | 5.1 | 5.1 | 10.2 | 2.5 | 2.6 | 5.1 |
| Gujarat | 0.6 | 1.4 | 2.0 | 29.3 | 64.9 | 94.2 | 1.5 | 3.2 | 4.7 | 0.7 | 1.6 | 2.4 |
| Delhi | 0.5 | 0.3 | 0.9 | 120.7 | 74.6 | 195.4 | 6.0 | 3.7 | 9.8 | 3.0 | 1.9 | 4.9 |
| Madhya Pradesh | 0.6 | 0.6 | 1.2 | 37.3 | 40.6 | 77.9 | 1.9 | 2.0 | 3.9 | 0.9 | 1.0 | 1.9 |
| Rajasthan | 3.6 | 3.6 | 7.2 | 60.1 | 58.9 | 119.0 | 3.0 | 2.9 | 6.0 | 1.5 | 1.5 | 3.0 |
| Uttar Pradesh | 2.0 | 5.5 | 7.5 | 33.5 | 90.0 | 123.5 | 1.7 | 4.5 | 6.2 | 0.8 | 2.3 | 3.1 |
| West Bengal | 1.6 | 0.7 | 2.3 | 80.6 | 35.9 | 116.4 | 4.0 | 1.8 | 5.8 | 2.0 | 0.9 | 2.9 |
| Andhra Pradesh | 0.5 | 1.3 | 1.8 | 44.1 | 114.5 | 158.5 | 2.2 | 5.7 | 7.9 | 1.1 | 2.9 | 4.0 |
| Bihar | 0.9 | 1.6 | 2.5 | 9.6 | 15.8 | 25.4 | 0.5 | 0.8 | 1.3 | 0.2 | 0.4 | 0.6 |
| Odisha | 4.1 | 1.6 | 5.7 | 42.2 | 16.3 | 58.5 | 2.1 | 0.8 | 2.9 | 1.1 | 0.4 | 1.5 |
| Karnataka | 4.3 | 11.8 | 16.1 | 105.1 | 290.1 | 395.2 | 5.3 | 14.5 | 19.8 | 2.6 | 7.3 | 9.9 |
| Jammu & Kashmir | 1.1 | 0.1 | 1.2 | 54.8 | 5.3 | 60.1 | 2.7 | 0.3 | 3.0 | 1.4 | 0.1 | 1.5 |
| Punjab | 2.3 | 5.4 | 7.7 | 59.6 | 143.1 | 202.7 | 3.0 | 7.2 | 10.1 | 1.5 | 3.6 | 5.1 |
| Telangana | 2.3 | 8.7 | 11.0 | 56.0 | 210.6 | 266.6 | 2.8 | 10.5 | 13.3 | 1.4 | 5.3 | 6.7 |
| Haryana | 2.3 | 5.1 | 7.4 | 38.7 | 85.6 | 124.3 | 1.9 | 4.3 | 6.2 | 1.0 | 2.1 | 3.1 |
| Chandigarh | 0.8 | 0.3 | 1.1 | 314.8 | 157.2 | 472.0 | 15.8 | 7.9 | 23.6 | 7.9 | 3.9 | 11.8 |
| Jharkhand | 1.5 | 2.1 | 3.6 | 28.4 | 41.4 | 69.8 | 1.4 | 2.1 | 3.5 | 0.7 | 1.0 | 1.7 |
| Kerala | 3.6 | 5.8 | 9.5 | 107.6 | 173.4 | 281.0 | 5.4 | 8.7 | 14.1 | 2.7 | 4.3 | 7.0 |
| Tripura | 3.9 | 0.2 | 4.1 | 109.8 | 5.9 | 115.7 | 5.5 | 0.3 | 5.8 | 2.8 | 0.1 | 2.9 |
| Rest of India | 3.4 | 1.9 | 5.3 | 70.6 | 42.2 | 112.8 | 3.5 | 2.1 | 5.6 | 1.8 | 1.1 | 2.8 |
| **India** | **1.9** | **3.2** | **5.1** | **53.0** | **87.9** | **140.9** | **2.6** | **4.4** | **7.0** | **1.3** | **2.2** | **3.5** |

Despite fewer hospitals, the number of hospital beds, ICU beds, and ventilators in Chandigarh is higher than the other States and UTs of India. However, the state like Bihar, Odisha, Jammu and Kashmir, Jharkhand, Madhya Pradesh, and Gujarat face a severe deficiency in health facilities. The Union health ministry reported on May 10, 2020; there are 7,740 dedicated Covid-19 facilities in 483 districts in the country including all states and central government facilities, to fight against the pandemic. Also, there are 6,56,769 isolation beds, 3,05,567 beds for confirmed cases, 3,51,204 beds for suspected cases, 99,492 oxygen supported beds, 1,696 facilities with oxygen manifolds, and 34,076 ICU beds (IANS, 2020). Though there are several quarantine centres have been created, many lack necessary infrastructure. The patients complained that there were only three washrooms and five large bedrooms for more than 40 people when they were quarantined in Delhi (Thacker 2020).

At present, Maharashtra (highest Covid-19 burden) has only 2.6 hospital, 188 hospital beds, 9.4 ICU beds, and 4.7 ventilators per lakh population. In Mumbai, about 30% of Covid-19 cases who are admitted as symptomatic, including 3% in need of critical care (Barnagarwala, 2020). Though this percentage is small, the city is still lacking hospital beds for these critical patients. The available facilities are divided into three layers, i.e., Dedicated Covid Hospitals (DCH), Dedicated Covid Health Centres (DCHCs), and Dedicated Covid Care Centre (DCCC). The DCH admits only critically ill patients who require ICU or ventilator support or patients who are gasping for breath whereas DCHCs admits moderately sick (cough, cold, and continued fever) Covid-19 patients and DCCC quarantine people who are high-risk suspects, slum-dwellers who cannot practice social distancing at home and asymptomatic positive cases, or cases with mild symptoms. However, there is plenty of space to admit people in DCHCs and DCCC, and the beds are limited for the infected Covid-19 patients in DCHs.

In this section, we focused on the availability of government hospital beds in other Covid-19 affected states. Using the data from National Health Profile-2019, it is observed that there are 20,172 government hospital beds in Gujarat (29.3 per one lakh population), 24,383 in Delhi (120.7 per one lakh population), 77,532 in Tamil Nadu (102 per one lakh population), and 47,054 in Rajasthan (60.1 per lakh population). The capacity of ICU beds is not sufficient in these states (Gujarat: 1009; Delhi: 1219; Tamil Nadu: 3877; and Rajasthan: 2353). An estimated 5-10% of total patients will require critical care in the form of ventilator support (Singh et al., 2020; Joshi, 2020). With the estimated number of 47,481 ventilators available in the country will be deficit in near future to cater to a growing number of active Covid-19 patients and the burden of other communicable and non-communicable diseases. Clearly, the growing demand for ventilators is going to outstrip the limited supply really soon.

Besides this, the health workers are instructed to wear the mandatory Personal Protective Equipment (PPE) to protect themselves from harmful biological agents or contaminated surfaces. Apart from this, the shortage of PPE for doctors, nurses, and other health workers are now becoming a global issue. India also lacks much needed PPE kit for those who screen, test, and treat Covid-19 patients. The nurses and health care workers were forced to use raincoats, scarves, motorcycle helmets to cover their faces in the absence of masks (Sarda, 2020). Apart from PPE kit, the government faced challenges with Covid-19 testing laboratories. Currently, India has only 569 testing laboratories (396 govt. and 173 private) against its 1.35 billion population (ICMR, 2020). The procurement of more Cobas 6800 testing machine, with the capacity to test around 1200 samples per day is essential.

## Discussion and Conclusion

Testing to detect the cases is one of the important strategies to deal with Covid 19 epidemic. To minimize the spread of the infection, it is vital to find out the detected cases, trace their contacts and quarantine infected persons. The number of tests per million populations may not provide accurate information about the adequacy of the tests rather the test positive rate provides a better measure. If we will say that the country or the state is in right direction, then the test positive rate should be low and it should decrease. States such as Maharashtra and Gujarat shows the test positive rate is increasing; it is almost double since last one month which is a concern. Therefore, the testing is grossly inadequate in these states. The test positive rate in India is almost stabilized around 4 percent since last one month. In order to bring the states and the country in right track, the test positive rate to bring down to below 2 percent as is observed from the successful example of Kerala,

During the lockdown, the epidemic curve of Covid 19 was flattered at somewhat desired level but the critical component of this post-lockdown strategy is adequate testing (Lancet, 2020). The immediate challenge is to keep infections at manageable levels and to test, trace contacts, isolate patients, and implement Covid care plans. The procurement of higher number of high throughput machine, the Cobas 6800 testing machine, is need of the hour. India must also pay much higher attention to the health sector and recognize the importance of having strong public sector capacity, especially in primary care and at the district level.

The health infrastructure of India made enormous progress over the past decades. The life expectancy has crossed 67 years, infant and under-five mortality rates are declining as is the rate of disease incidence. The diseases, such as polio, yaws, and guinea worm disease, etc. have been eradicated. In spite of the progress, communicable diseases are expected to continue, and the non-communicable diseases are now leading cause of death due to the absence of adequate health facilitates at the primary, secondary, and tertiary levels (Narain, 2016). At present, India faces the triple burden of diseases- the infectious diseases, the challenge of non-communicable diseases (NCDs), and the emerging of Covid-19 pandemics. The existing health infrastructure in India is already over-stretched and needs to be strengthening to face these challenges in the twenty-first century. The life after Covid-19 will not be the same. The current disaster reiterated the fact that healthcare and life sciences are the biggest opportunities for our country. According to Global health experts, India does not have the proper infrastructure or financial capability to deal with a massive public health disaster (Changoiwala, 2020). There are also severe shortages of medical staff and supplies throughout India. This pandemic is a lesson and wake-up call for the government worldwide to change in priorities in GDP and needs to be prepared to deal with the same crisis in the future. There is a need to increase the public health spending in India at least upto the global average of 6 percent, which should focus both preventive and curative care. In the current situation, there is an urgent need to make a sizeable allocation to the annual health financing in India. The priority should be the creation of more laboratories and hospitals, storage of PPE kit, testing kit, and indigenously developed vaccines. Besides, spending on research and training is another priority area. The government needs to address the gap in medical advancements (i.e., latest technology, medical knowledge) and research to provide better care giving at all levels. Looking at the severity of Covid-19, there is a need to scale-up public health services, the number of beds and physicians, medical equipment, medicines, and care packages for public health emergencies. Public-private partnership is another priority area, especially when the country is in a public health emergency. The government needs to strengthen medical infrastructure through the public-private partnership to make India not only the ‘Pharmacy of the World’ but also the ‘Laboratory of the World’.

## Data Availability

The paper has used data from publicly available secondary sources. https://api.Covid19india.org/csv/

## Acknowledgement

Authors would like to thank Mr. Sushanta Mandal for his contribution in preparing the map.

## References

Banerjee, C. (2020). Why parts of India can’t ramp up Covid-19 testing, retrieved from https://timesofindia.indiatimes.com/india/why-parts-of-india-cant-ramp-up-Covid-19-testing/articleshow/75080738.cms

Barnagarwala, T. (2020). Nature of the crisis: Why Mumbai is running out of beds for critical Covid patients, retrieved from https://indianexpress.com/article/explained/mumbai-coronavirus-Covid-19-cases-hospital-beds-deaths-6413025/C:\Users\Admin\Downloads\ on 17th May 2020.

Basu, D. (2020a), Indian States must increase COVID-19 testing. But by How Much? Retrieved from https://science.thewire.in/health/states-coronavirus-testing/

Basu, J. (2020b). COVID-19: Bengal has highest death rate, least testing in India, retrieved from https://www.downtoearth.org.in/news/governance/Covid-19-bengal-has-highest-death-rate-least-testing-in-india-70728.

Cascella, M., Rajnik, M., Cuomo, A., Dulebohn, S. C., & Di Napoli, R. (2020). Features, evaluation, and treatment coronavirus (COVID-19).In Statpearls [internet].StatPearls Publishing.

Changoiwala P. (2020). Covid-19 Threatens to Overwhelm India’s Health Care System, retrieved from https://undark.org/2020/04/14/Covid-19-india/.

Chauhan, C. & Kawoosa, V. (2020). Covid-19 crisis: At 32 per million, India lags far behind on testing, retrieved from https://www.hindustantimes.com/india-news/some-of-india-s-biggest-states-have-abysmal-testing-numbers/storv-8CoLZEmBG6NxKc0Z3BI9KI.html.

Gao, J., Tian, Z., & Yang, X. (2020). Breakthrough: Chloroquine phosphate has shown apparent efficacy in treatment of COVID-19 associated pneumonia in clinical studies. Bioscience trends, 14(1): 72–73.

IANS (2020). Adequate health infrastructure, facilities for COVID-19: Govt., retrieved from https://health.economictimes.indiatimes.com/news/industry/adequate-health-infrastructure-facilities-for-Covid-19-govt/75660316

Indian Council of Medial Research (2020). Total Operational (initiated independent testing) Laboratories reporting to ICMR. Department of Health Research. Ministry of Health and Family Welfare, retrieved from: https://www.icmr.gov.in/pdf/Covid/labs/COVID_Testing_Labs_20052020.pdf.

Joshi, M. (2020).91 Covid patients in ICUs, 27 on ventilators in Delhi, retrieved from https://indianexpress.com/article/cities/delhi/91-Covid-patients-in-icus-27-on-ventilators-in-delhi-6403762/.

Kannan, S., Ali, P. S. S., Sheeza, A., & Hemalatha, K. (2020).COVID-19 (Novel Coronavirus 2019)-recent trends. European review for medical and pharmacological sciences, 24(4), 2006–2011.

Kapoor et al. (2019). COVID-19 in India: State-wise estimates of current hospital beds, intensive care unit (ICU) beds and ventilators, Washington D.C.: The Centre for Disease Dynamics, Economics and Policy.

Lancet, T. (2020).India under COVID-19 lockdown.Lancet (London, England), 395(10233), 1315, retrieved from https://www.thelancet.com/action/showPdf?pii=S0140-6736%2820%2930938-7

Nair, S. (2020). Coronavirus Hotspots in India: Complete list of 130 COVID-19 hotspot districts, all metro cities red zones, retrieved from https://www.jagranjosh.com/current-affairs/coronavirus-hotspot-areas-in-india-what-are-hotspots-know-all-Covid-hotspots-1586411869-1

Narain, J. P. (2016). Public health challenges in India: seizing the opportunities. Indian journal of community medicine: official publication of Indian Association of Preventive & Social Medicine, 41(2): 85–88.

National Health Profile 2019 (Issue No. 14).Central Bureau of Health Intelligence. Ministry of Health and Family Welfare, GoI, retrieved from http://www.cbhidghs.nic.in/showfile.php?lid=1147

Remuzzi, A., & Remuzzi, G. (2020).COVID-19 and Italy: what next?. The Lancet, 395(10231): 1225–1228, retrieved from https://www.sciencedirect.com/science/article/pii/S0140673620306279

Registrar General of India. (2019). Population Projection for India and States 2011–2036. New Delhi, retrieved from https://nhm.gov.in/New_Updates_2018/Report_Population_Projection_2019.pdf

Sarda, P. (2020). India’s PPE crisis puts workers in the line of fire, retrieved from https://www.forbesindia.com/article/coronavirus/indias-ppe-crisis-puts-workers-in-the-line-of-fire/59073/1

Sharma, M. (2020). Health security must get the attention it deserves in India’s response to Covid19, retrieved from: https://www.orfonline.org/expert-speak/health-security-must-get-the-attention-it-deserves-in-indias-response-to-Covid19-65010/

Singh, A. K., Singh, A., Shaikh, A., Singh, R., & Misra, A. (2020). Chloroquine and hydroxychloroquine in the treatment of COVID-19 with or without diabetes: A systematic search and a narrative review with a special reference to India and other developing countries. Diabetes & Metabolic Syndrome: Clinical Research & Reviews, 14(3): 241–246.

Singh, et. al., (2020). COVID-19: Is India’s health infrastructure equipped to handle an epidemic?, retrieved from https://www.brookings.edu/blog/up-front/2020/03/24/is-indias-health-infrastructure-equipped-to-handle-an-epidemic/

Singhal, T. (2020). A review of coronavirus disease-2019 (COVID-19). The Indian Journal of Pediatrics, 87: 281–286.

Thacker, T. (2020).Covid-19 spread: Poor condition of quarantine facilities come into focus in India, retrieved from https://economictimes.indiatimes.com/news/politics-and-nation/poor-conditions-of-quarantine-facilities-come-into-focus/articleshow/74738682.cms?from=mdr

Thakur et al,. (2020). Delay in test results has Centre, states worried, retrieved from https://www.hindustantimes.com/india-news/delay-in-test-results-has-centre-states-worried/story-VJU6Doa3RSyxmWsWSf073J.html

United Nations Development Programme (UNDP). COVID-19 pandemic: Humanity needs leadership and solidarity to defeat the coronavirus, retrieved from https://www.undp.org/content/undp/en/home/coronavirus.html

Unnithan, P. (2020). Kerala confirmed first novel coronavirus case in India, retrieved from https://www.indiatoday.in/india/story/kerala-reports-first-confirmed-novel-coronavirus-case-in-india-1641593-2020-01-30

Velavan, T. P., & Meyer, C. G. (2020).The COVID 19 epidemic. Tropical medicine & international health, 25(3): 278–280.

